# A SARS-CoV-2 Label-Free Surrogate Virus Neutralization Test and a Longitudinal Study of Antibody Characteristics in COVID-19 Patients

**DOI:** 10.1101/2021.01.19.21250137

**Authors:** Yiqi Ruben Luo, Cassandra Yun, Indrani Chakraborty, Alan H.B. Wu, Kara L. Lynch

**Affiliations:** Department of Laboratory Medicine, University of California San Francisco and Zuckerberg San Francisco General Hospital, San Francisco, CA, USA; Gator Bio, Palo Alto, CA, USA

**Keywords:** SARS-CoV-2, label-free technology, neutralizing antibody titer, longitudinal study

## Abstract

**Background:** The laboratory-based methods to measure the SARS-CoV-2 humoral response include virus neutralization tests (VNTs) to determine antibody neutralization potency. For ease of use and universal applicability, surrogate virus neutralization tests (sVNTs) based on antibody-mediated blockage of molecular interactions have been proposed.

**Methods:** A surrogate virus neutralization test established on a label-free immunoassay platform (LF-sVNT). The LF-sVNT analyzes the binding ability of RBD to ACE2 after neutralizing RBD with antibodies in serum.

**Results:** The LF-sVNT neutralizing antibody titers (IC50) were determined from serum samples (n=246) from COVID-19 patients (n=113), as well as the IgG concentrations and the IgG avidity indices. Although there is variability in the kinetics of the IgG concentrations and neutralizing antibody titers between individuals, there is an initial rise, plateau and then in some cases a gradual decline at later timepoints after 40 days post-symptom onset. The IgG avidity indices, in the same cases, plateau after the initial rise and did not show a decline.

**Conclusions:** The LF-sVNT can be a valuable tool in clinical laboratories for the assessment of the presence of neutralizing antibodies to COVID-19. This study is the first to provide longitudinal neutralizing antibody titers beyond 200 days post-symptom onset. Despite the decline of IgG concentration and neutralizing antibody titer, IgG avidity index increases, reaches a plateau and then remains constant up to 8 months post-infection. The decline of antibody neutralization potency can be attributed to the reduction in antibody quantity rather than the deterioration of antibody avidity, a measure of antibody quality.

**Summary:** A surrogate virus neutralization test established on a label-free immunoassay platform (LF-sVNT). Using the LF-sVNT and other assays, 246 serum samples from 113 COVID-19 patients were measured. We observed the time course of antibody characteristics beyond 200 days post-symptom onset.

## Introduction

Since the start of the COVID-19 pandemic in early 2020, much research has focused on the kinetics and magnitude of the immune response and measurable correlates of acquired immunity. SARS-CoV-2 infection can be detected indirectly by measuring the host immune response. Most immunocompetent individuals with symptomatic infections develop detectable SARS-CoV-2 antibodies within 2 weeks of symptom onset (1–3). The SARS-CoV-2 IgG antibody response is more robust in severe cases of COVID-19 at all time-points after seroconversion (1). However, the antibody neutralization potency, depicted as neutralizing antibody titers, cannot be directly obtained from the antibody concentrations (2). Intraindividual variation in the quantity of antibodies produced indicates that serological responses may not be equivalent in terms of future protection. Early reports suggest that the SARS-CoV-2 IgG concentrations can wane over time, however, it is unclear if the antibodies that persist are capable of neutralizing the virus (4). While the antibody quantity may decline, the quality of remaining IgG antibodies, as determined by the measurement of antibody avidity, or functional affinity, increases over time post-symptom onset (5). It is still unknown how antibody production following vaccination will compare to that of acquired disease. Further studies are needed to determine how durable the humoral immune response is following acquired disease and vaccination.

Laboratory-based methods used to measure the SARS-CoV-2 humoral response include qualitative and quantitative methods for total antibody or antibody subclasses (IgG, IgM, IgA) (1,4), IgG avidity (5), and antibody neutralization potency. Plaque reduction neutralizing tests, also known as conventional virus neutralization tests (cVNTs), measure SARS-CoV-2 neutralizing antibody titer and involve the use of live pathogens and target cells (6,7). Experiments using pandemic pathogens like SARS-CoV-2 impose special safety requirements and cannot be implemented in most clinical laboratories, limiting the widespread availability of testing. Methods using pseudovirus (pVNTs) have been published, however, these can take days to obtain results (8–10). For ease of use and universal applicability, surrogate virus neutralization tests (sVNTs) based on antibody-mediated blockage of molecular interactions have been proposed (10–15). An sVNT measures the competitive inhibition of the interaction between a viral structural protein and angiotensin-converting enzyme 2 (ACE2), since ACE2 is the receptor of SARS-CoV-2 on host cells (16). Thus, sVNTs can be designed with compatibility for routine clinical laboratory settings. Like cVNTs, sVNTs detect neutralizing antibodies in an isotype-independent manner, offering a key advantage over antibody concentration assays. The SARS-CoV-2 spike protein receptor-binding domain (RBD) is the favored choice as the viral structural protein used in sVNTs because (1) it is the binding domain located on the spike protein responsible for viral entry into host cells (17); (2) it has better binding characteristics in comparison to spike protein S1 subunit and nucleocapsid protein (11); and (3) it is a highly specific target for antibodies and has less cross-reactive epitopes with other coronavirus (18,19).

Recently, open-access label-free technologies have emerged as a novel solution for next-generation immunoassays in clinical laboratories (20,21). These technologies can measure the time course of immunoreactions in real-time without attaching a reporter (enzyme, fluorophore, etc.); thus it provides fast measurement, simple operation with automation, and ease of assay development and troubleshooting. One such technology, thin-film interferometry (TFI), has been used for the routine therapeutic drug monitoring of the monoclonal antibody therapeutics and associated anti-drug antibodies in human serum. The same technology has been applied to the measurement of SARS-CoV-2 IgG avidity (5). This paper describes the development and validation of a novel label-free surrogate virus neutralization test (LF-sVNT) using the TFI technology. The method was used to measure neutralizing antibodies in a cohort of COVID-19 patients and determine if they correlated with total SARS-CoV-2 IgG concentration or avidity. Serial serum samples collected from mild to severe COVID-19 patients were measured out to 8 months post-symptom onset to determine the kinetics and durability of the neutralizing antibody response.

## Materials and Methods

### Ethical review

All serum samples used in the analysis were remnant specimens obtained following routine clinical laboratory testing. The study protocol was approved by the Institutional Review Board of the University of California San Francisco. The committee deemed that written consent was not required for use of remnant specimens.

### Specimens and reagents

Individual and serial serum samples (n=246) from patients diagnosed with COVID-19 (PCR-confirmed) (n=113) were obtained for testing. The patients were 63% male and 75% Hispanic, with a median age of 51 years. Fifty-eight patients (51%) were hospitalized and 55 patients (49%) were outpatients. Of the hospitalized patients, 33 (57%) were admitted to the intensive care unit (ICU), 25 (43%) received mechanical ventilation, and 2 died while in the ICU. The sampling time span of the patients ranged from 5 to 225 days after symptom onset.

Recombinant RBD and ACE2 were purchased from Sino Biological (Wayne, PA). A human monoclonal anti-RBD IgG1 antibody was obtained from Absolute Antibody (Oxford, UK), and a goat anti-human IgG antibody (anti-IgG) from Jackson Immunoresearch (West Grove, PA). The TFI label-free immunoassay analyzer and the sensing probes coated with RBD were manufactured by Gator Bio (Palo Alto, CA). The Pylon 3D fluorescence immunoassay analyzer was manufactured by ET Healthcare (Palo Alto, CA).

### Label-free surrogate neutralization assay

A label-free surrogate neutralization assay (LF-sVNT) was established on the TFI label-free immunoassay analyzer to measure neutralizing antibody titers in serum samples. The LF-sVNT analyzes the binding ability of RBD to ACE2 after neutralizing RBD with antibodies in serum. The sensing probes in use were pre-coated with RBD. Each serum sample was diluted in a series (1:50, 1:100, 1:250, 1:500, 1:1000, 1:2000) in running buffer (PBS at pH 7.4 with 0.02% Tween 20, 0.2% BSA, and 0.05% NaN_3_) for analysis. The LF-sVNT was carried out by dipping a sensing probe sequentially into a sample and reagents. The LF-sVNT protocol consisted of two cycles: the first cycle included the steps of 1) dipping the sensing probe in running buffer for a baseline, 2) forming RBD-Ab immune complexes with a sample, 3) rinsing the sensing probe in running buffer, 4) forming RBD-ACE2 immune complex with 12 μg/ml ACE2 in running buffer; the second cycle included only the first and last steps of the first cycle. Between the two cycles, the sensing probe was regenerated in 10 mM glycine at pH 2.0 to strip off the molecules bound to RBD. The concentration of ACE2 was set at 12 μg/ml (0.14 μM), several times higher than the reported affinity constant between RBD and ACE2 monomer 18.5 nM (22), to ensure the saturation of RBD on the sensing probe. The first cycle measured the binding ability of RBD to ACE2 after neutralization, and the second cycle provided the full binding ability of RBD without neutralization. In each cycle, the recorded time course of signals, as known as the sensorgram, was recorded. The readout measured the signal increase in the step of forming RBD-ACE2 immune complex, representing the quantity of the immune complex on the sensing probe. The neutralization index was calculated as the ratio of the readout in the first cycle to that in the second cycle, presented as a percentage, meaning the residual binding ability of RBD to ACE2 after neutralization. The illustration of the assay protocol and example sensorgrams are shown in Figure 1.

**Figure 1.**
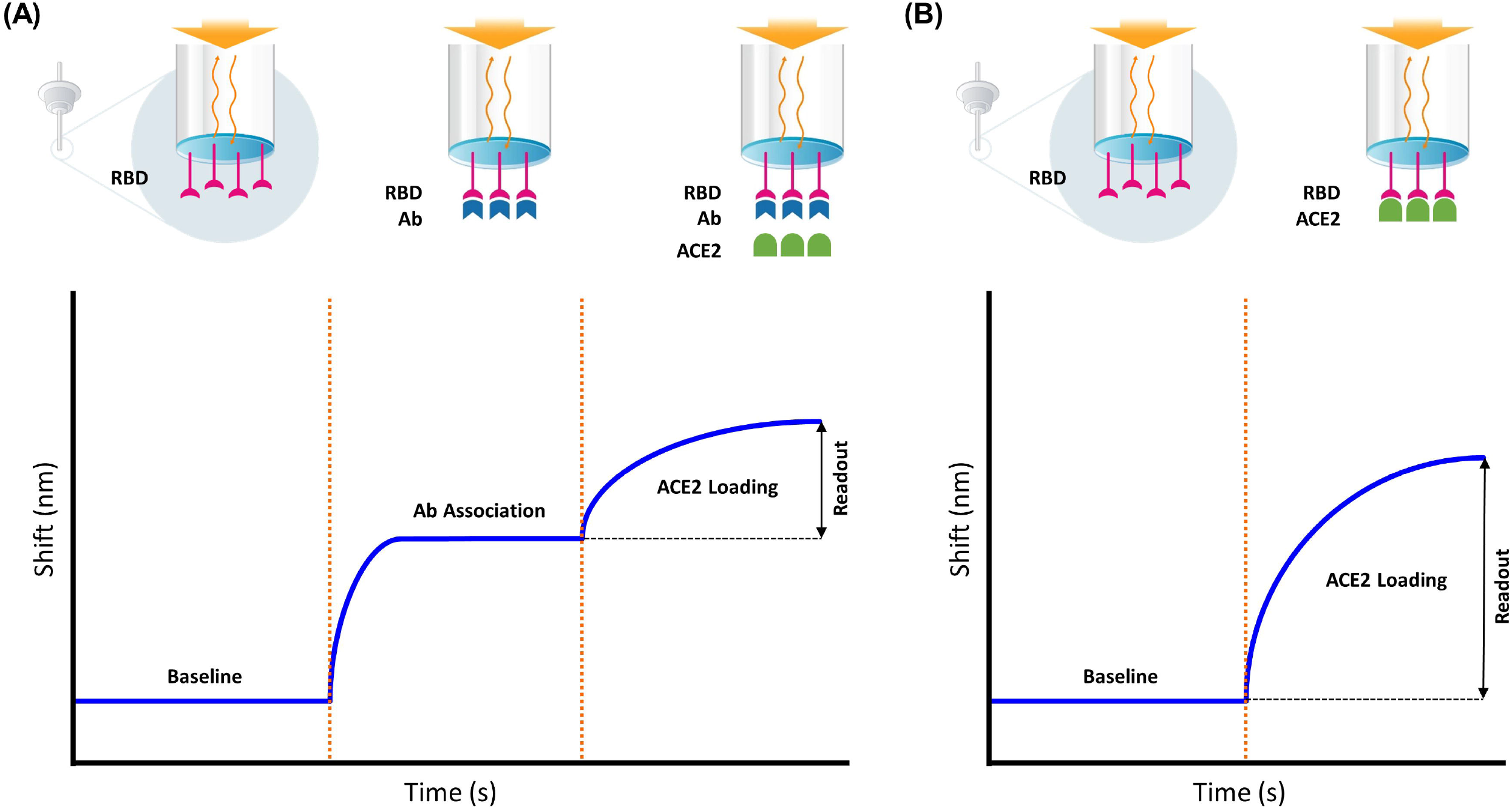
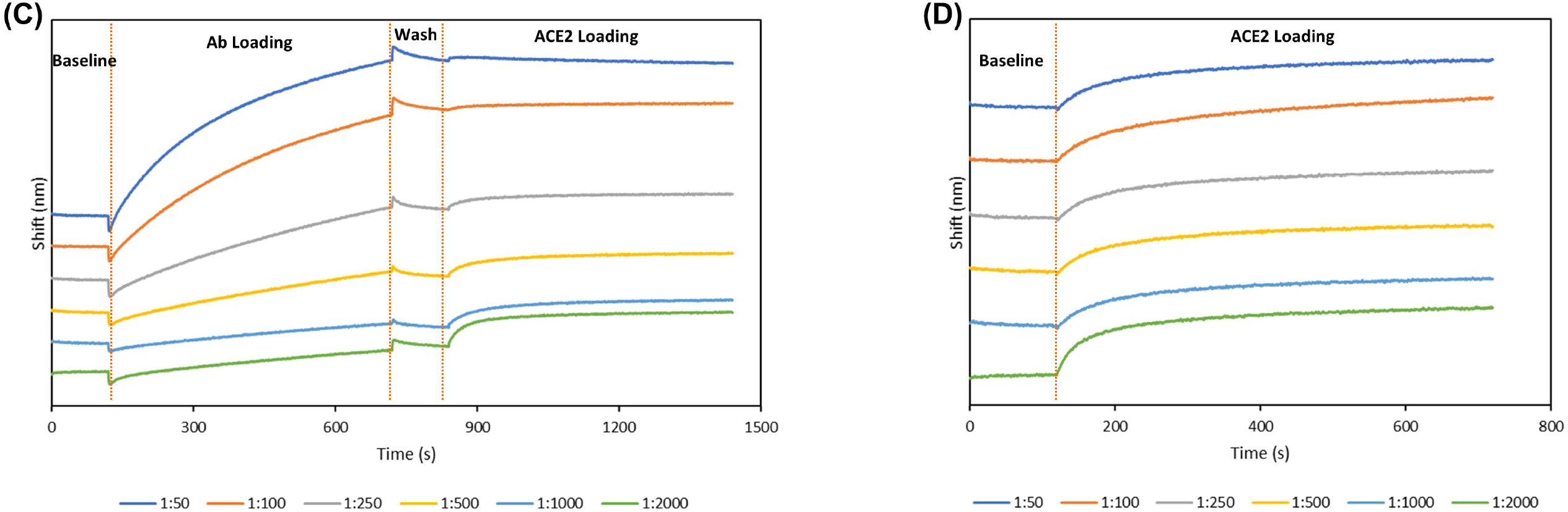
Illustration of the LF-sVNT protocol and example sensorgrams. A) the first cycle measuring the binding ability of RBD to ACE2 after neutralization; B) the second cycle measuring the full binding ability of RBD without neutralization; C) the first-cycle sensorgrams and D) and second-cycle sensorgrams of a dilution series of one serum sample (1:50, 1:100, 1:250, 1:500, 1:1000, 1:2000).

The precision of the LF-sVNT was verified using a spiked serum sample, i.e. a human monoclonal SARS-CoV-2 anti-RBD IgG at 20 μg/ml in a negative serum sample. The spiked serum sample was measured by 5 sensing probes, and the measurement was repeated 3 times using the same sensing probes with regeneration in between. When measuring patient serum samples, the spiked serum sample was used as a positive control, and the results of the positive control were used to calculate the precision across different batches. Specificity was evaluated using a set of 22 serum samples from individuals that tested RT-PCR negative for SARS-CoV-2 and positive for other respiratory viruses (4 coronavirus HKV1, 1 coronavirus 229E, 2 coronavirus OC43, 5 human rhinovirus/enterovirus, 4 human metapneumovirus, 3 respiratory syncytial virus, 2 parainfluenza type 1 virus, and 1 adenovirus).

To obtain the neutralizing antibody titer (IC50) for each serum sample, the neutralization indices were plotted against dilutions, and the points were fitted using a linear interpolation model. The reciprocal of the dilution resulting in a 50% neutralization index was defined as the neutralizing antibody titer.

### Label-Free IgG avidity assay and fluorescence IgG concentration assay

A label-free IgG avidity assay was also established using the TFI technology, as reported previously (5). The sensing probes and running buffer were the same as those in the LF-sVNT. Each serum sample was 10-fold diluted in running buffer for analysis. The IgG avidity assay protocol included the steps of 1) dipping the sensing probe in running buffer for a baseline, 1) forming RBD-IgG immune complex on the sensing probe, 2) dissociating loosely bound IgG using either running buffer or 3 M urea in running buffer, and 3) forming RBD-IgG-Anti-IgG immune complex using 10 μg/ml anti-IgG in running buffer. The signal increase in the final step, which is proportional to the quantity of RBD-IgG-Anti-IgG immune complex on the sensing probe, was measured. The IgG avidity index was calculated as the ratio of the readout with the dissociation agent (urea) to the reference (running buffer), presented as a percentage. A fluorescence IgG concentration assay was carried out on the Pylon 3D fluorescence immunoassay analyzer, as reported previously (1).

### Conventional virus neutralization test

A cVNT was used for method comparison with the LF-sVNT in a subset of serum samples spanning the range of the assay. The experiment was implemented at Colorado State University. A 2-fold dilution series of each serum sample was prepared in Hank’s balanced salt media (BA-1). Each dilution was mixed with an equal volume of SARS-CoV-2 virus suspension and incubated for 60 min at 37°C with 5% CO_2_. The mixture was then added to a Vero cell suspension and incubated for 45 min. After incubation, a first layer of overlay (2X MEM with 1% agarose) was added to each well, and the plates were again placed in the incubator. After 24 hours, a second overlay with 0.05% neutral red dye was added, and the plates were incubated at 37°C. Viral plaques were counted the following day and the 50% cutoff was calculated based on negative control plaque counts. The PRNT50 titer reported is the reciprocal of the highest dilution of serum that inhibits ≥50% of the plaques relative to the control.

## Results

The precision (% coefficient of variation) of the neutralization indices for dilutions 1:50, 1:100, 1:250, 1:500, 1:1000, 1:2000 across 5 sensing probes was 9.0%, 2.2%, 2.9%, 4.4%, 3.0%, 1.7%, and across 3 repeats (same sensing probes with regeneration in between) was 7.5%, 2.9%, 5.9%, 6.0%, 4.2%, 1.8%, respectively. The precision of the neutralization indices across different batches (n=8) was 9.8%, 4.7%, 9.5%, 8.3%, 3.8%, 2.3% for the same dilutions, respectively. Serum sample from COVID-19 negative and other respiratory virus positives patients were all negative (IC50<50).

The LF-sVNT neutralizing antibody titers (IC50) were determined from serum samples (n=246) from COVID-19 patients (n=113), as well as the IgG concentrations and the IgG avidity indices. For correlation analyses, only one sample per patient per week was included (n=190). The neutralizing antibody titers showed a weak correlation with IgG concentrations (Figure 2A, least squares linear regression correlation coefficient 0.72). There was no correlation between neutralizing antibody titers and IgG avidity (Figure 2B). LF-sVNT was compared to a cVNT, in a subset of serum samples selected to cover the range of LF-sVNT neutralizing antibody titers (IC50 <50 to 2000) (Figure 2C, least squares linear regression correlation coefficient 0.64). The method for calculating titers between the two methods was different, liner interpolation for LF-sVNT and the highest dilution that inhibited 50% of plaque formation for cVNT, limiting the direct comparison. High LF-sVNT neutralizing antibody titers (≥1000) predominately appeared between 10 and 40 days after disease onset and were higher in critically ill patients (Figure 2D). The neutralizing antibody titers begin to decline after 40 days post-symptom onset out to 225 days (the latest timepoint measured).

**Figure 2.**
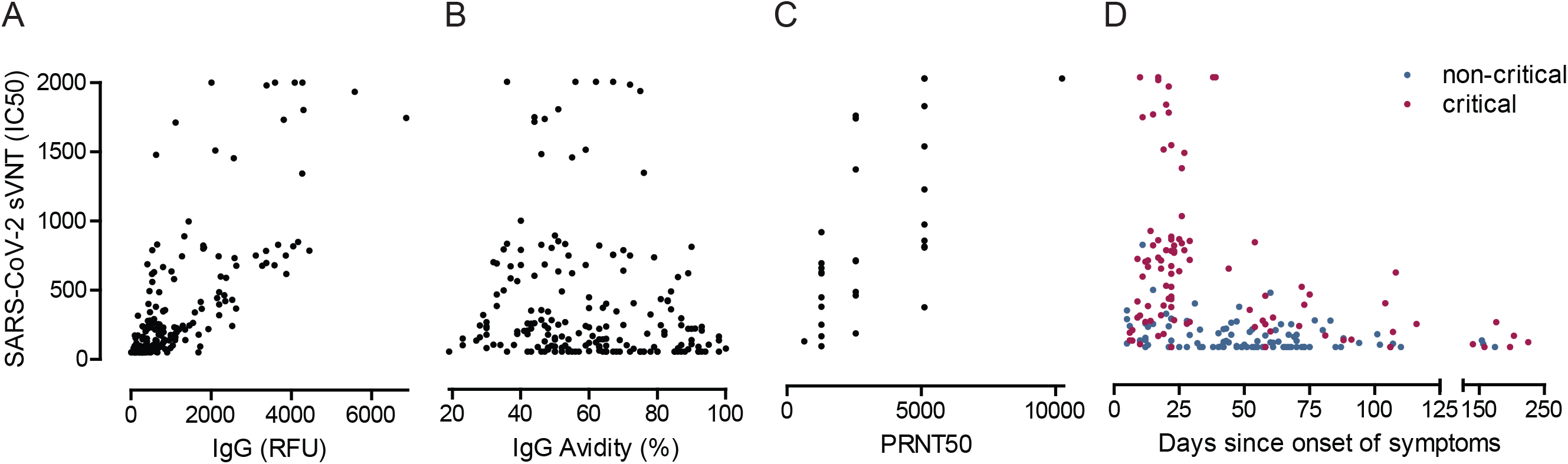
Comparison of LF-sVNT neutralizing antibody titers with, A) IgG concentrations and B) IgG avidity indices (n=190). C) Comparison of LF-sVNT and the cVNT neutralizing antibody titers obtained from 30 serum samples. D) LF-sVNT neutralizing antibody titers plotted against days since onset of symptoms. All titers <50 (n=39) were plotted at 50 and titers >2000 (n=4) were plotted at 2000. For A, B and C only one sample was included per subject per week.

To further characterize the apparent decline in LF-sVNT neutralizing antibody titers and the correlation with IgG concentrations and avidity, all available paired serum samples from week 4 and 3-8 months post-symptom onset were measured (Figure 3). For 20 paired samples, IgG concentrations and LF-sVNT neutralizing antibody titers declined in all but 2 and 1 subjects, respectively. In contrast, the SARS-Cov-2 IgG avidity increased in all but 1 subject. For 9 subjects, 4 or more serial samples with at least one sample beyond 30 days post-symptom onset were available for measuring the kinetics of the antibody response (Figure 4). Although there is variability in the kinetics of the IgG concentrations and neutralizing antibody titers between individuals, there is an initial rise, plateau and then in some cases a gradual decline at later timepoints after 40 days post-symptom onset. The IgG avidity indices, in the same cases, plateau after the initial rise and did not show a decline.

**Figure 3.**
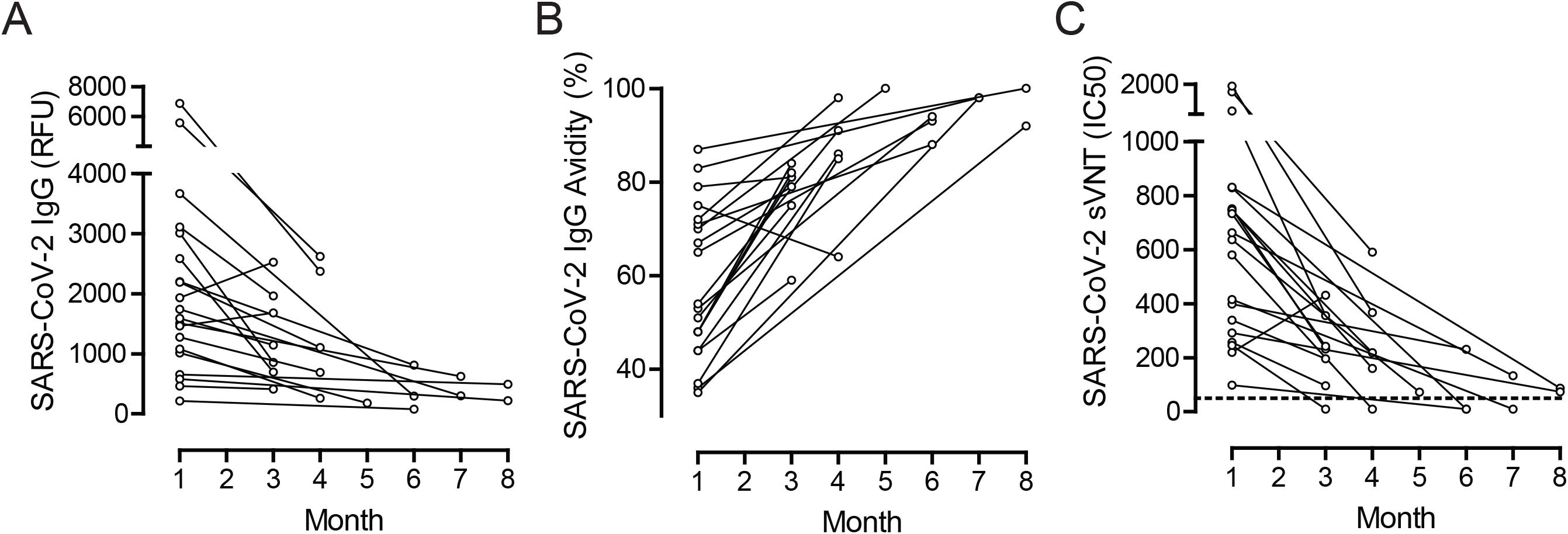
IgG concentration (A), IgG avidity index (B), and LF-sVNT neutralizing antibody titer (C) of paired serum samples from week 4 and 3-8 months post-symptom onset (n=20). The horizontal dashed line indicates the limit of detection for the LF-sVNT.

**Figure 4.**
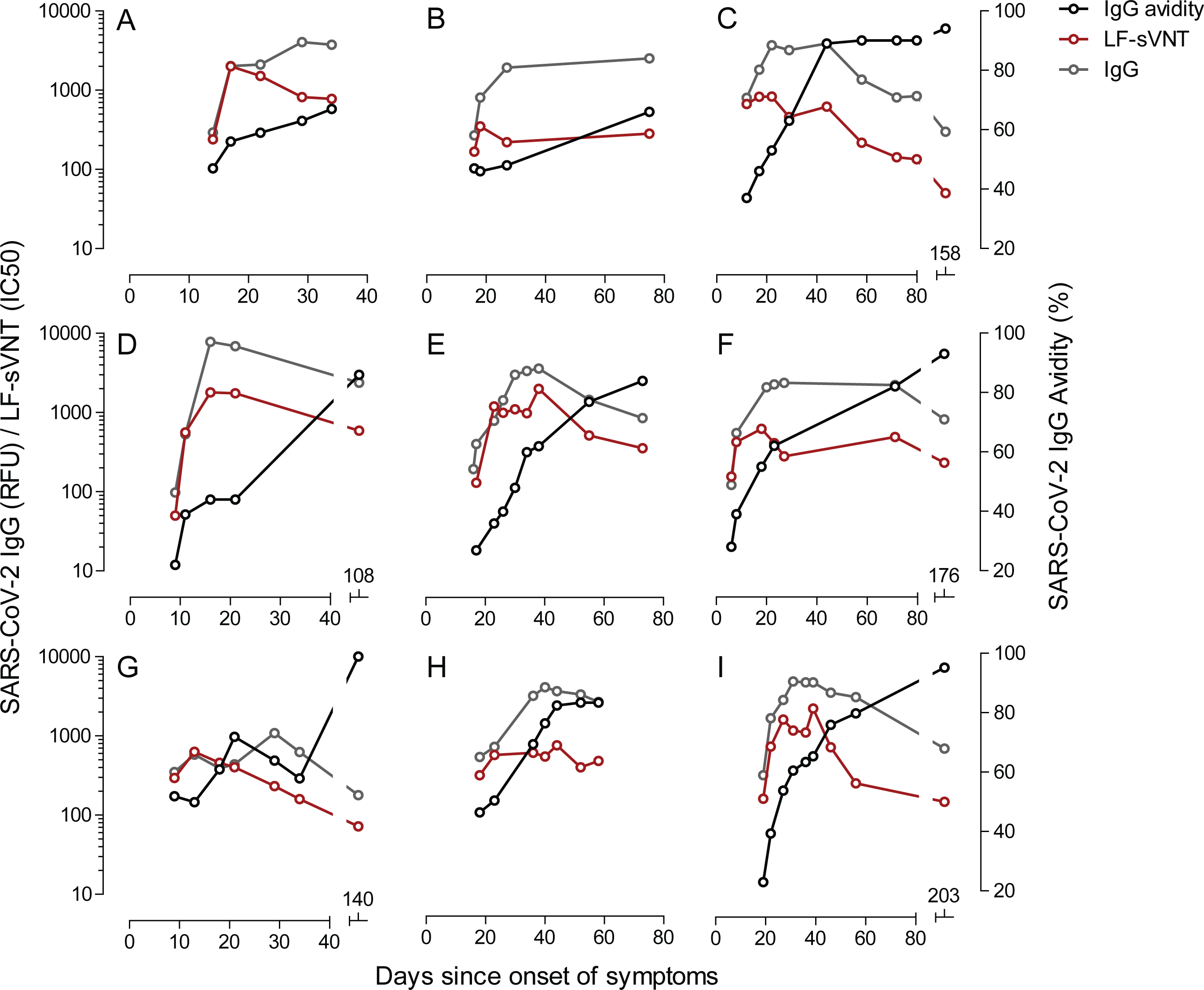
Kinetics of LF-sVNT neutralizing antibody titer, IgG concentration, and IgG avidity for 9 patients by days after symptom onset. All cases with ≥ 4 serial time points and enough remaining sample volume to perform all tests were included.

## Discussion

The clinical utility of serological testing for SARS-CoV-2 continues to be a subject of debate. To date, multiple qualitative and quantitative methods for SARS-CoV-2 total antibodies, antibody subclasses, antibody avidity and neutralization potency, have been developed and used to characterize the humoral response to active infection through convalescence. Despite the advancing knowledge from these studies, questions remain regarding lasting protection following infection and vaccination. A method that can rapidly provide data on SARS-CoV-2 antibody neutralization potency and is amenable to a high-throughput clinical laboratory setting may offer a diagnostic test to accurately determine protective immunity. This remains to be determined. Here we present an sVNT method using a novel label free technology, that correlated with absolute IgG antibody concentration and a cVNT, and can be performed with a rapid turn-around time. The LF-sVNT employed a sensing probe coated with RBD to mimic the surface of SARS-CoV-2 and ACE2 as an equivalent of host cells. Although the RBD-ACE2 interaction mainly characterizes viral attachment (17,23), it mimics the virus-host cell interaction since viral attachment is the main determinate of viral entry into host cells.

The label-free technology employed allows for real-time monitoring of the RBD-ACE2 interaction and antibody mediated blockage. Thus, in comparison with other sVNTs, the LF-sVNT removes the steps of reporter attachment (if not pre-conjugated) and signal generation, e.g. attaching a secondary antibody-labeled enzyme (10), attaching a streptavidin-labeled enzyme or fluorophore (14,15), and carrying out a color-generating enzymatic reaction (10–14). This advantage of LF-sVNT eliminates any possible interference to the viral protein-ACE2 complex during these steps and decreases testing time compared to other published sVNT methods (10,11,14). The LF-sVNT provides process efficiency for fully automated, random-access testing, compared to batch testing for most other sVNTs. The real-time monitoring of the RBD-ACE2 interaction can be used for quality assurance to reduce experimental error rate. The LF-sVNT can be easily modified to incorporate mutated variants of RBD. Thus, the LF-sVNT can be a valuable tool in clinical laboratories for the assessment of the presence of neutralizing antibodies to COVID-19.

Recent reports provide evidence for a decline in SARS-CoV-2 neutralizing antibody titers in patients over 40 days after disease onset and the measurement was carried out with samples collected up to 94 days (4). Further assessment of antibody neutralization potency for a longer time frame is still necessary to determine the longevity of the neutralizing antibody response. In this study, the overall decline of neutralizing antibody titer was consistent with previous reports, however, this data is the first to provide longitudinal neutralizing antibody titers beyond 200 days post-symptom onset (24–28). As shown in Figure 4, for the analysis of paired specimens, with the first specimen 4 weeks post-symptom onset and the second 3-8 months post-symptom onset, in 17 out of 20 patients the neutralizing antibody titers declined over 60% within 8 months, and in 5 patients the titers dropped to an undetectable level (IC50 <50). Similarly, IgG concentration declined over 60% within 8 months for 10 out of 20 patients. However, whether or not a decay in neutralizing antibody response increases the risk for reinfection remains unanswered (29).

Despite the decline of IgG concentration and neutralization antibody titer, IgG avidity index increases, reaches a plateau and then remains constant up to 8 months post-infection. Prior to this study, longitudinal data on SARS-CoV-2 IgG avidity only extended to 90 days post-symptom onset (5). The decline of antibody neutralization potency can be attributed to the reduction in antibody quantity rather than the deterioration of antibody avidity, a measure of antibody quality. Whether the maintenance of antibody quality over time correlates with protection to SARS-CoV-2 infection remains unknown, however, it is a positive indicator of prolonged humoral immunity, which might relate to the continuous presence of SARS-CoV-2 memory B cells (30). As anamnestic immune responses against SARS-CoV-2 rechallenge have been demonstrated in rhesus macaques (31), this observation enhances the expectations of longevity of immune protection after infection or vaccination. On the other hand, the need for repeat vaccination or booster for the current mRNA COVID-19 vaccines has not been established beyond the two-dose primary series (32). The longitudinal study of antibody neutralization potency and IgG avidity can aid in the assessment of the need for continual revaccination and the required frequency.

## Data Availability

None

## Acknowledgment

The authors thank Dr. Raymond Goodrich and Lindsay Hartson at the Infectious Disease Research Center, Colorado State University for carrying out the cVNT.

